# Evaluating GPT-4 as a Clinical Decision Support Tool in Ischemic Stroke Management

**DOI:** 10.1101/2024.01.18.24301409

**Authors:** Amit Haim, Mark Katson, Michal Cohen-Shelly, Shlomi Peretz, Dvir Aran, Shahar Shelly

## Abstract

Cerebrovascular diseases are the second most common cause of death worldwide and one of the major causes of disability burden. Advancements in artificial intelligence (AI) have the potential to revolutionize healthcare delivery, particularly in critical decision-making scenarios such as ischemic stroke management. This study evaluates the effectiveness of GPT-4 in providing clinical decision support for emergency room neurologists by comparing its recommendations with expert opinions and real-world treatment outcomes. A cohort of 100 consecutive patients with acute stroke symptoms was retrospectively reviewed. The data used for decision making included patients’ history, clinical evaluation, imaging studies results, and other relevant details. Each case was independently presented to GPT-4, which provided a scaled recommendation (1-7) regarding the appropriateness of treatment, the use of tissue plasminogen activator (tPA), and the need for endovascular thrombectomy (EVT). Additionally, GPT-4 estimated the 90-day mortality probability for each patient and elucidated its reasoning for each recommendation. The recommendations were then compared with those of a stroke specialist and actual treatment decision. The agreement of GPT-4’s recommendations with the expert opinion yielded an Area Under the Curve (AUC) of 0.85 [95% CI: 0.77-0.93], and with real-world treatment decisions, an AUC of 0.80 [0.69-0.91]. In terms of mortality prediction, out of 13 patients who died within 90 days, GPT-4 accurately identified 10 within its top 25 high-risk predictions (AUC = 0.89 [95% CI: 0.8077-0.9739]; HR: 6.98 [95% CI: 2.88-16.9]), surpassing supervised machine-learning models. This study demonstrates the potential of GPT-4 as a viable clinical decision support tool in the management of ischemic stroke. Its ability to provide explainable recommendations without requiring structured data input aligns well with the routine workflows of treating physicians. Future studies should focus on prospective validations and exploring the integration of such AI tools into clinical practice.

## Introduction

The advent of GPT-4, launched by OpenAI in March 2023, marked a significant milestone in the evolution of artificial intelligence (AI) and its applications in various domains, including healthcare. GPT-4, a model under the umbrella of Generative Pretrained Transformers (GPT), exemplifies the advancement in large language model (LLM) technology.^1,2^ The foundational architecture of this technology involves training on extensive datasets, enabling the model to function as a ‘few-shot learner’. This capability allows GPT-4 to adapt to new domains and continuously refine its performance through ongoing learning.^1,3^

In the realm of clinical medicine, the potential applications of LLMs like GPT-4 are particularly intriguing. These models offer promise as supportive tools for healthcare professionals, aiding in the efficient summarization of patient data, assisting in decision-making processes, and potentially improving the accuracy and speed of medical interventions.^4,5^ Recent research has underscored the capabilities of GPT-4 in complex medical tasks. Notably, the model has demonstrated proficiency in examinations akin to the United States Medical Licensing Examination (USMLE), achieving scores that meet or nearly meet the passing thresholds.^6^ Additionally, in assessments modeled after neurology board exam questions, GPT-4 has shown a high accuracy rate, improving with repeated attempts.^7,8^

The management of acute ischemic stroke (AIS) presents a critical and time-sensitive challenge in clinical settings. The approach to diagnosing and treating AIS requires a synthesis of information including patient symptoms, physical and neurological examinations, medical history, and imaging results. Despite the availability of established guidelines by the American Heart Association/American Stroke Association (AHA/ASA) for stroke management,^9-11^ the pivotal role of the treating physician’s judgment remains. Variability in clinical presentations and the urgent need for decision-making underscore the potential value of AI-assisted tools in this context. Moreover, predicting early mortality in AIS is essential for guiding treatment decisions, optimizing resource allocation in healthcare settings, facilitating effective communication with patients and their families, supporting research and clinical trials, and contributing to quality improvement initiatives. In accordance, several traditional machine-learning models have been trained for this task in recent years.^12-14^

Here, we leveraged patient data from the emergency department of a large referral hospital, focusing on individuals presenting with stroke symptoms, to evaluate the effectiveness of GPT-4 in delivering accurate clinical decisions for the treatment of AIS. We also assessed its proficiency in predicting 90-day mortality outcomes. We aimed to quantify the extent to which an advanced language model like GPT-4 can augment the clinical decision-making process, potentially contributing to improved patient outcomes in one of the most critical areas of emergency medicine.

## Results

### Patient demographics and clinical data

We generated a cohort from 100 consecutive cases of patients presenting with acute stroke symptoms at the emergency department of Rambam Healthcare Campus. All cases underwent full clinical and radiological evaluation in the emergency setting for acute stroke and were fully evaluated by a neurologist (**Table 1**; **Figure 1A**). Revascularization treatment was administered to 78 of the patients: 36 were treated with tPA, 30 with EVT, and 12 received both. Within this cohort, 13 patients died within 90 days and 21 in total. Overall, 17 cases were classified as ‘complex’ when not fitting exact treatment guidelines.^9^ The data for each case encompassed demographics, NIHSS^15^ scores, timing of arrival to brain CT, onset of symptoms, and details from textual brain imaging results and risk factors that were available as medical history at the time of admission to the ER (**Supplementary Table 1**).

**Table 1.** Study cohort clinical information and demographics.

| **Variable** | **Simple cases** | **Complex cases** |
| --- | --- | --- |
| **All** | 83 | 17 |
| **Females (%)** | 38 (45.78) | 7 (41.17) |
| **Age, Median (range)** | 75 (72) | 71 (44) |
| **First NIHSS (Median, range)** | 12 (25) | 5 (19) |
| **Median Time to CT, hours (range)** | 1.83 (3.40) | 4.45 (3.42) |
| **Brain CT findings (%)** |  |  |
| **LVO** | 48 (57.83) | 7 (41.17) |
| **MCA** | 47 (56.62) | 4 (23.52) |
| **PCA** | 8 (9.63) | 4 (23.52) |
| **Risk Factors** |  |  |
| **HTN (%)** | 51 (61.44) | 10 (58.82) |
| **DM** | 35 (42.16) | 3 (17.64) |
| **Dyslipidemia** | 36 (43.37) | 6 (35.29) |
| **Smoking** | 11 (13.25) | 4 (23.52) |
| **CKD** | 11 (13.25) | 0 (0) |
| **Obese** | 5 (6.02) | 0 (0) |
| **Cancer** | 9 (10.84) | 1 (5.88) |
| **HF** | 7 (8.43) | 1 (5.88) |
| **Cardiac Arrhythmia** | 19 (22.89) | 2 (11.76) |
| **Family history for CAD** | 1 (1.2) | 0 (0) |
| **TPA (%)** | 29 (34.93) | 7 (41.17) |
| **EVT (%)** | 29 (34.93) | 1 (5.88) |
| **TPA + EVT (%)** | 12 (14.45) | 0 (0) |
| **90-day mortality (%)** | 11 (13.25) | 2 (11.76) |
| **Overall Mortality (%)** | 17 (20.48) | 4 (23.52) |

**Figure 1.**
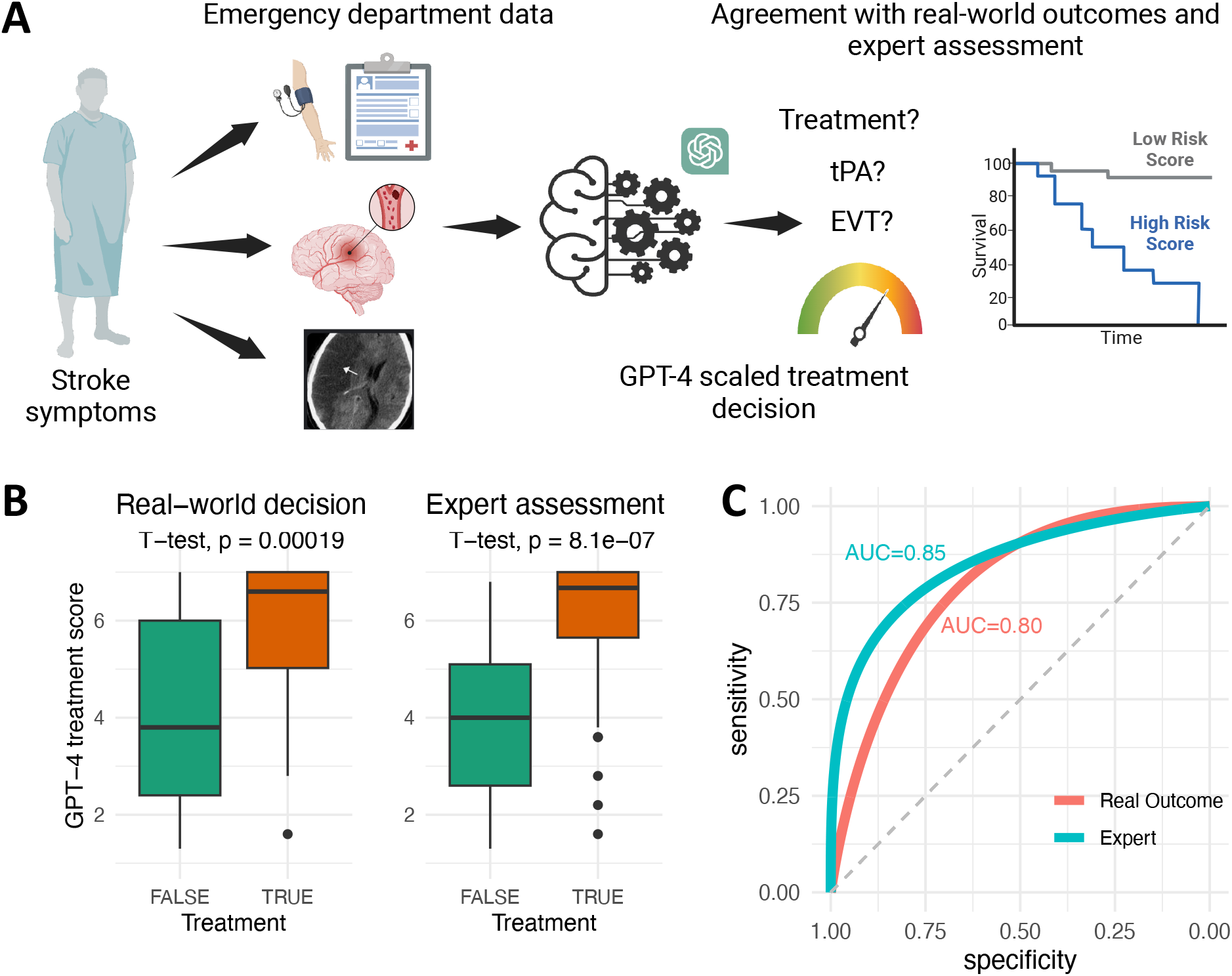
Study Design and GPT-4 Performance Evaluation. **A**. Illustration of the study design involving 100 consecutive stroke patients who underwent a comprehensive stroke workup, including perfusion, angiography, and non-contrast brain CT upon arrival at the Emergency Room. Clinical information, demographics, comorbidities, and CT perfusion results were recorded. The textual reports from these investigations were entered into the GPT-4 API, which was instructed to provide scores indicating whether to treat the patient, whether to administer tPA, whether to pursue EVT, and an estimate of 90-days mortality (Created with BioRender.com). **B**. Boxplots presenting average scores of GPT-4 assessments for decision to treat (y-axis). The comparison is made against real-world decisions and expert assessments of each case (TRUE – to treat the patient, FALSE – to not treat). **C**. ROC curves and AUC scores of GPT-4 average scores for decision to treat, compared to real-world decisions and expert assessments.

A stroke specialist, blinded to the outcomes, retrospectively reviewed each case. In 82 of the cases, the expert’s decisions aligned with the actual treatments administered. Of note, the expert recommended against treating 11 patients who received treatment and suggested treatment for 7 who did not receive any. Concerning specific treatments, full agreement was observed in 61 cases, although the expert more frequently recommended combining tPA and EVT than what was observed in practice (Cohen’s Kappa = 0.51, signifying moderate agreement).

### GPT-4 clinical decisions

Independently, each case was assessed with GPT-4, generating a treatment recommendation scale from 1 (intervention not recommended) to 7 (highly recommended) (**Figure 1A**; **Supplementary Table 2**). To account for the variability in GPT-4 responses, each case was assessed 5 times. Cohen’s Kappa for treatment scores across runs ranged from 0.56 to 0.73. As expected, the pre-defined ‘complex’ cases demonstrated significantly greater variance between runs (p-value = 0.02).

Comparing GPT-4’s treatment scale to both the expert’s decision and the actual treatment revealed that the average scores from GPT-4 for patients who were treated were, on average, 1.9 points higher than those not treated (p-value < 0.001), and 2.1-point difference in comparison to the expert decision (p-value < 0.001; **Figure 1B**). The average scores provided an area under the ROC curve (AUC-ROC) of 0.80 [95% CI: 0.69-0.91] compared to real-world decisions, and 0.85 [95% CI: 0.77-0.93] compared to the expert decision (**Figure 1C**). These average scores for AUCs were higher than those of each independent run (**Supplementary Figure 1**). Additionally, removing the clinical presentation narrative from GPT-4’s analysis resulted in a drop in AUC to 0.70 with real-world decisions and 0.72 with the expert (**Supplementary Figure 1**), highlighting the importance of unstructured narrative data in treatment decision-making. Similarly, setting the temperature of GPT-4 to 0 resulted in AUC of 0.70 and 0.72 with the real-world and the expert, respectively, suggesting the need to allow GPT-4 more creativity to obtain better decisions.

Using a score threshold of 4, we observed 22 disagreements between GPT-4 and real-world treatment and 20 disagreements with the expert decision. Notably, a significant proportion of these disagreements coincided with cases where the expert and real-world decisions diverged, with 18 out of 30 such cases showing this dual disagreement. Moreover, complex cases were more prone to discrepancies, as 7 disagreements with real-world and 5 with the expert were noted among the 17 complex cases. The specialist examined the explanatory text produced by GPT-4 for all discrepancies between the model and their blinded assessments, evaluating whether they agreed that the explanatory text, as part of the original model output, was logical and could be deemed good practice. Of the 20 instances where disagreements occurred, in three cases the expert, after having carefully considered GPT-4’s detailed explanations, conceded that GPT-4’s assessment was preferable to their original decision. In additional two cases the expert acknowledged that GPT-4’s suggested approach was indeed acceptable and aligned with viable treatment options. In instances where the expert disagreed with GPT-4’s reasoning, the disagreements primarily revolved around three key issues. Firstly, GPT-4 inaccurately associated abnormal angiographic findings with clinical presentations. An illustrative case is that of a patient with stenosis of the right-sided middle cerebral artery (MCA) who presented with right hemiparesis (case 94). Despite these two elements potentially being anatomically unrelated, GPT-4 linked them erroneously. The second notable issue pertained to ethical considerations, particularly in a case involving a patient with active laryngeal cancer and cognitive decline. According to guidelines, the patient was deemed eligible for treatment, but the expert’s decision was to not proceed with treatment as life expectancy was short and he was palliative (case 14). Thirdly, discrepancies arose in deviations from guidelines, particularly in cases of distal thrombectomies. For instance, in the case of an over 90 year-old patient with M2 obstruction (considered distal thrombus), GPT-4 recommended against treatment, which is the established guidelines, however, the expert call was to proceed with thrombectomy due to high NIHSS score and good results in such cases in the past from personal experience (case 54).

In assessing GPT-4’s ability to choose the best treatment option, it showed near-perfect agreement with real-world decisions in recommending EVT: GPT-4 suggested EVT for all patients (42 of the 42) treated with EVT (average score >4). The expert suggested EVT for 55 patients, of which 50 were also recommended EVT by GPT-4, corresponding to an AUC of 0.94 [95% CI: 0.89-0.98] with real-world decisions and 0.95 [95% CI: 0.90-0.99] with the expert (**Figure 2A**). For tPA treatment, GPT-4 recommended it for 38 of the 48 patients who received it, showing a closer agreement with the expert. Of the 41 patients recommended for tPA by the expert, GPT-4 agreed on 35, corresponding to an AUC of 0.77 [95% CI: 0.68-0.86] with real-world decisions and 0.82 [95% CI: 0.73-0.90] with the expert (**Figure 2B**).

**Figure 2.**
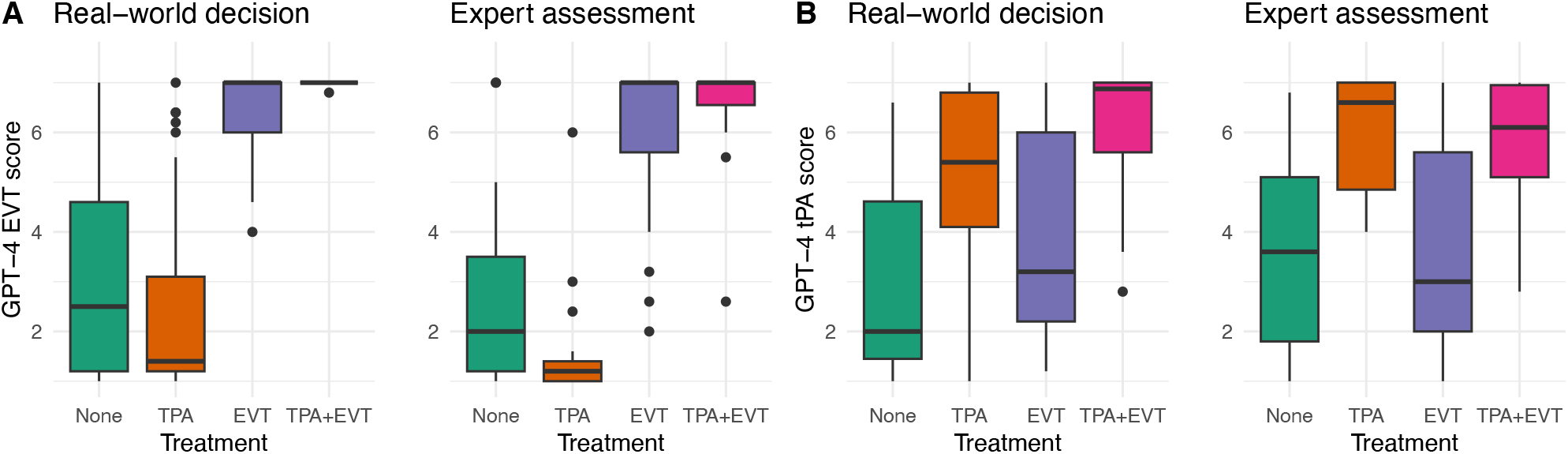
GPT-4 treatment type scores. **A**. Boxplots depict GPT-4 treatment type scores, with the Y-axis representing probability score (1-7 scale). Each treatment category is color-coded: green for no intervention, orange for tPA (Tissue Plasminogen Activator), purple for Endovascular treatment (EVT), and pink for tPA and EVT. **A**. GPT-4 scores for EVT, stratified by real-world decisions and expert assessments. **B**. GPT-4 scores for tPA, stratified by real-world decisions and expert assessments.

### Mortality risk

We further evaluated the ability of GPT-4 to predict 90-day mortality. The model estimated an average mortality risk of 55.1% for patients who died within 90 days, compared to 31.5% for survivors (p-value < 0.001), yielding an AUC of 0.89 [95% CI: 0.8077-0.9739] (**Figure 3A**). To contextualize these results, we compared GPT-4’s performance with that of two recent machine-learning models specifically trained for 90-day mortality prediction. In our cohort, the PRACTICE model^13^ achieved an AUC of 0.70, significantly worse than the GPT-4 predictions (log-rank p-value = 0.02), while the PREMISE model^14^ reached an AUC of 0.77 (p-value = 0.07) (**Figure 3A**). These comparisons underscore GPT-4’s remarkable accuracy in mortality risk assessment, outperforming specialized, trained predictive models.

**Figure 3.**
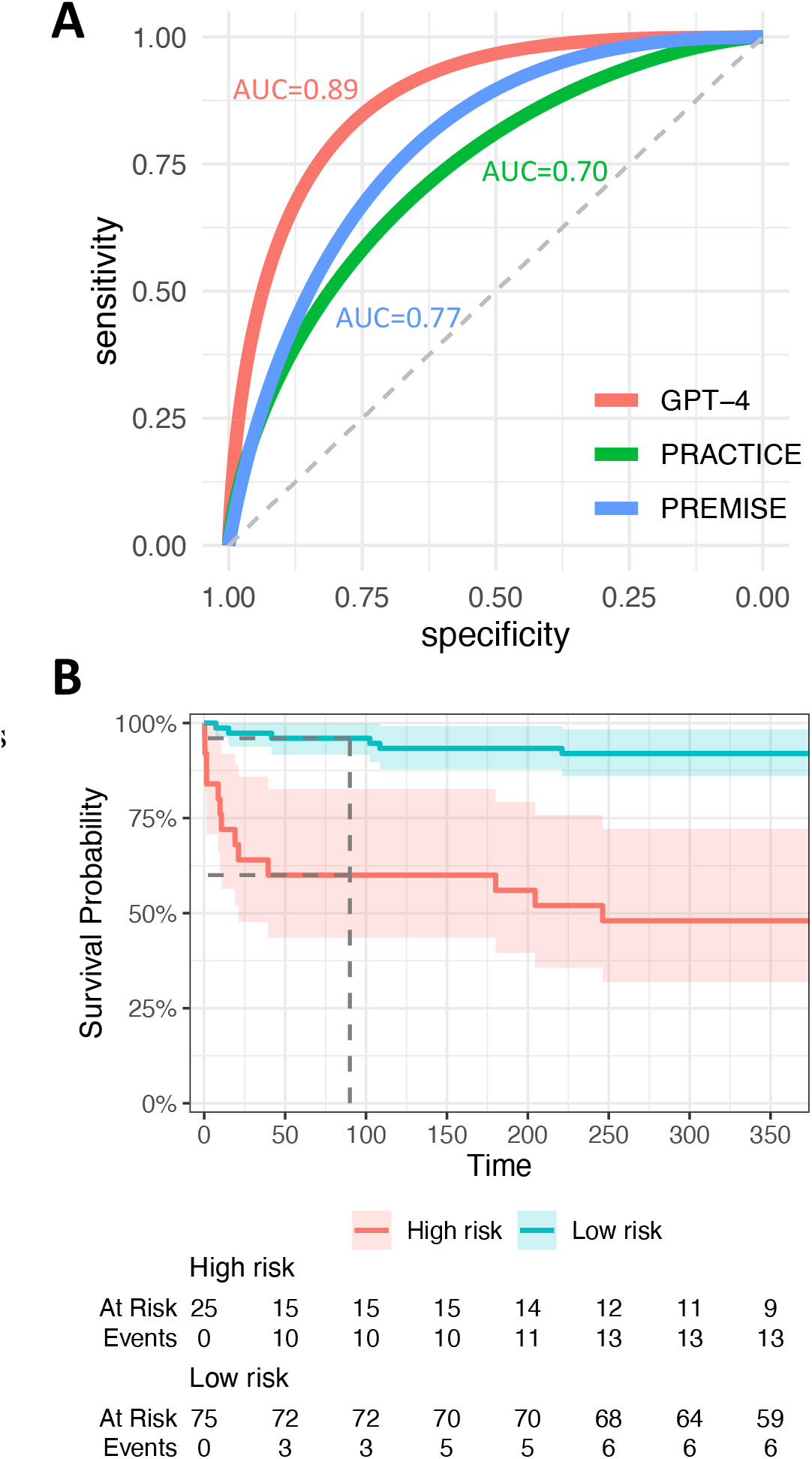
GPT-4 Mortality Predictions. **A**. ROC curve for 90-day mortality estimations by GPT-4 (red), PRACTICE (green), and PREMISE (blue). **B**. Kaplan-Meier plot stratifying individuals into low and high-risk categories for mortality based on GPT-4’s 90-day mortality estimations.

For identifying high-risk patients, we set a threshold at the top 25% of the cohort, which corresponded to a predicted mortality risk cut-off of 41%. Within this high-risk group, 10 patients passed away within 90 days of admission, and an additional 3 within the subsequent year (**Figure 3B**). Conversely, among the remaining 75 patients categorized as lower risk, only 3 deaths occurred within the 90-day period, and 6 in total during the first year. The calculated Hazard Ratio was 6.98 [95% CI 2.88-16.9; p-value <0.001], reinforcing the model’s capability in stratifying patients based on their mortality risk effectively.

## Discussion

This study introduces a pioneering application of a LLM predictive model, specifically GPT 4, to address acute ischemic stroke, the second most common cause of mortality and major cause of disability.^17-19^ The urgency of stroke care is particularly magnified in regions where access to specialized stroke units or even qualified physicians is limited, especially in countryside areas.^20,21^ The time-sensitive nature of stroke prognosis underscores the critical need for swift and accurate decision-making in these under-resourced healthcare facilities. The utility of GPT-4 in clinical practice is highlighted by its ability to operate seamlessly within existing treatment routines.^4^ The model relies solely on routine chart information available in emergency settings, making it particularly valuable in regions with limited access to neurology experts or areas with high patient volume and admission rates necessitate quick triage. This accessibility could democratize high-level medical consultation, extending expert-level decision-making to under-resourced healthcare facilities.

In our study, we also utilized GPT-4 to assess the predictive capacity for 90-day mortality in patients undergoing endovascular treatment for acute ischemic stroke. The GPT-4 model, utilizing a diverse range of clinical and imaging variables, demonstrated high accuracy in estimating mortality risk. Notably, the variables considered by GPT-4 encompass a wider spectrum, including clinical and imaging factors, offering a more comprehensive approach compared to existing models such as older scores such as HIAT and HIAT2, and newer scores such as PREMISE and PRACTICE. ^13,14,22,23^ Traditionally, healthcare predictive models rely heavily on collecting vast amounts of structured data and training specific machine learning algorithms. Contrarily, GPT-4 breaks this mold by providing comprehensive treatment recommendations and mortality risk based on narrative text, which is complicated to model in traditional machine-learning models. Our analyses highlighted the significance of unstructured data, as evidenced by the drop in prediction accuracy when the narrative clinical presentation was excluded. While traditional machine learning models struggle to process and interpret unstructured text, GPT-4 does so with apparent ease, showcasing its capability to handle complex medical data in a way that is more aligned with the natural flow of clinical information.

A crucial aspect of GPT-4’s application in healthcare is its explainability. Deep learning models often face challenges in providing clear reasoning for their decisions, a significant barrier in clinical practice where understanding the rationale behind a recommendation is crucial. Our analysis of the decision rationale provided by GPT-4 has further demonstrated its ability to effectively highlight key clinical considerations. The expert’s review confirmed that GPT-4’s explanations were not only correct but also offered valuable insights, thereby underscoring the model’s potential as an instrumental aid in clinical decision-making. This level of explainability enhances trust in AI-assisted decision-making and could pave the way for broader acceptance and integration of AI tools in medical settings.

Despite its promising results, our study has several limitations. We must acknowledge certain challenges in applying GPT-4, especially regarding its ability to assess ethical issues. The model may face difficulties in addressing the nuanced and complex ethical considerations intrinsic to medical decision-making. This limitation emphasizes the necessity for cautious and supplementary human oversight when deploying AI tools like GPT-4 in sensitive healthcare contexts. The occurrence of ‘hallucinations’ or erroneous outputs is another concern, although we demonstrated that running multiple assessments can mitigate this risk. Future research should focus on refining these methods to further reduce inaccuracies. Another consideration is the generalizability of these findings. Our study was conducted in a single center with a specific patient population. Further studies across diverse settings and larger populations are necessary to validate the efficacy and applicability of GPT-4 in various clinical environments.

In conclusion, our study introduces a groundbreaking approach to clinical decision support in stroke management using GPT-4. This model has shown the potential to process narrative text, provide explainable recommendations and enhance medical decision-making. As we continue to explore and refine this technology, it holds the promise of transforming patient care and improving outcomes in one of the most critical areas of medicine.

## Methods

### Ethical approval

Institutional review board/ethics committee approved this retrospective study in accidence of guidelines.

### Cohort Selection

This retrospective study comprised 100 consecutive cases from the emergency department of Rambam Healthcare Campus. All patients treated between January 2022 and April 2023 received a confirmed diagnosis of acute ischemic stroke. The inclusion criteria encompassed patients older than 18, an NIHSS^15^ score of 5 or higher(with the exception of patients 93 who received tPA off site), and less than 5 hours from symptom onset to undergoing a non-contrast CT of the brain. All included patients underwent non-contrast brain CT, CT angiography, and CT perfusion while in the ER. This cohort was specifically chosen for its alignment with AHA guidelines for acute stroke management^9^, making each patient a potential candidate for both tPA and EVT treatment. Seventeen patients, not meeting these criteria, were categorized as “complex” cases which the clinical scenario warranted extra consideration of off-guideline treatment options, and there was a need to assess the individual patient’s unique characteristics, medical history, and condition. For every patient, comprehensive medical records from their ER arrival, including imaging results, were collected, and translated from Hebrew to English. Exclusion criteria were patients with incomplete clinical data or where stroke was not the final diagnosis.

Clinical data for each patient included demographics, medical history, chief complaints, symptom onset time, physical and neurological examinations, NIHSS score, imaging results (including ASPECTS^16^ when available), treatment received, and mortality data. An experienced stoke specialist, blind to the outcomes, reviewed the cases and made treatment decisions among no treatment, tPA, EVT, or a combination of tPA and EVT. All data was deidentified, removing identifiers, names and dates.

### Analysis Pipeline

The analysis utilized the OpenAI API ‘create chat completion’ method with the model gpt-4-1106-preview. Default parameters were set (temperature = 1, top_p = 1, n = 1), and submissions were made using the R wrapper library ‘openai’.

The full prompt given to GPT-4 was as follows:

*“Imagine you are a board-certified neurologist in the emergency room. You are receiving a clinical case. Describe the best neurological approach leading to the best neurological outcome, and the lowest chance of mortality. Base your decision on the current guidelines and reason your decision. Note that in some cases patients should not be treated although the best treatment option due to the patient fragility*.

*Here is your case: <<CASE>>*

*First, provide the full reasoning. Next, based on your response, answer the questions below. Only return a number, no additional reasoning. Provide the results in a structured format as following: [A, B,C,D], where A is answer for q1, B for q2, C for q3 and D for q4. For example, an answer could be [4,3,2,40]*.

1. *Any intervention (tPA or EVT)? Answer with scale 1 to 7, where 1 is intervention not recommended and 7 is intervention is highly recommended*.
2. *Thrombolytic therapy (tissue plasminogen activator; tPA)? Answer with scale 1 to 7, where 1 is tPA not recommended and 7 is tPA is the best option*.
3. *Endovascular thrombectomy (EVT)? Answer with scale 1 to 7, where 1 is EVT not recommended and 7 is EVT is the best option*.
4. *What is your estimation for 90-day mortality probability? Provide estimation even if there is not enough information. Use the scale 0 and 100*.*”*

To assess the reliability of GPT-4 responses, each case underwent five submissions, we well as an additional submission without the accompanying clinical presentation narrative. For every treatment decision, GPT-4 provided a narrative explanation. In 95% of cases, GPT-4 returned responses in the requested structure, which were automatically scraped with R. Unstructured responses were manually entered. For estimations provided as a range, the average was used. If GPT-4 provided a number with a greater symbol (e.g., >50), the number was recorded with an additional 5. In 0.8% of cases, GPT-4 did not return numeric responses for treatment decisions, and in 8.6% of responses, it did not provide a 90-day mortality estimate.

### Statistical Analysis

GPT-4’s responses were scaled from 1 to 7 for treatment decisions and from 0 to 100 for 90-day mortality estimations. Averages were calculated across the five repeats. All statistical analyses were conducted using R version 4.3.2, employing base R functions, pROC 1.18.5, and survival 3.5.7. ROC curves were smoothed. Agreement between treatment decisions was measured using a linear weighted Cohen’s kappa coefficient, utilizing the psych 2.3.12 library.

## Data Availability

All data that was used in this study is available as a supplementary table.

## Acknowledgment and Author contributions

Study concept and design: SS & DA

Acquisition of data: SS, AM & DA

Analysis and interpretation: DA, SS, MK & SP

Draft of manuscript: DA & SS

Critical revision of the manuscript: AM, MK, SP & MCS

---

**Supplementary Figure 1.**
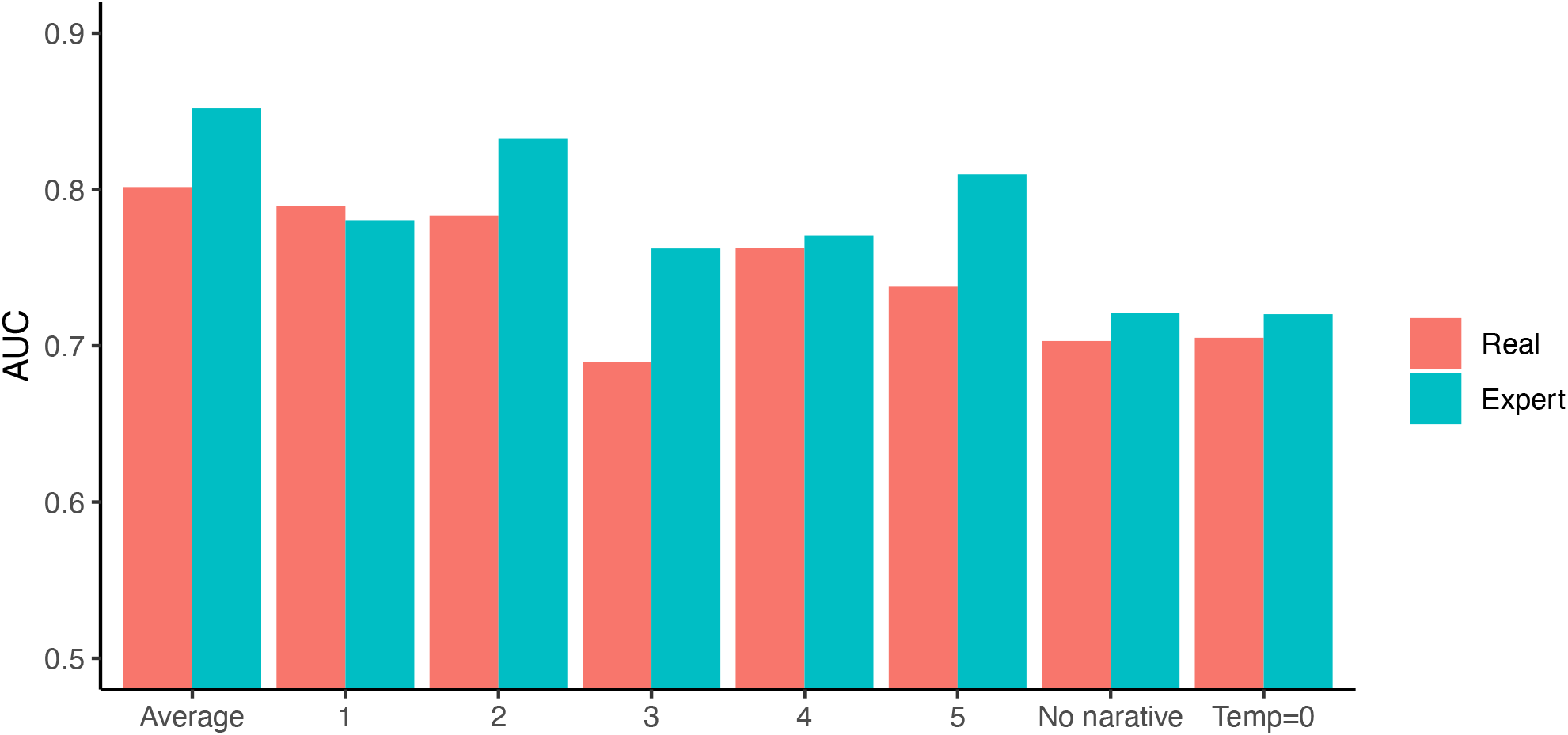
GPT-4 Assessments Performance. Area under the curve (AUC) for GPT-4 decision to treatment scores of each of the individual submissions (1-5) and the average. Each individual submission is lower than the average. In addition, we submitted the cases without the clinical presentation narrative, which yielded lower AUC (no narrative). Similarly, lower AUC was observed when cases were submitted with temperature=0.

