## Supplementary Figure 1 for "Evaluating GPT-4 as a Clinical Decision Support Tool in Ischemic Stroke Management"

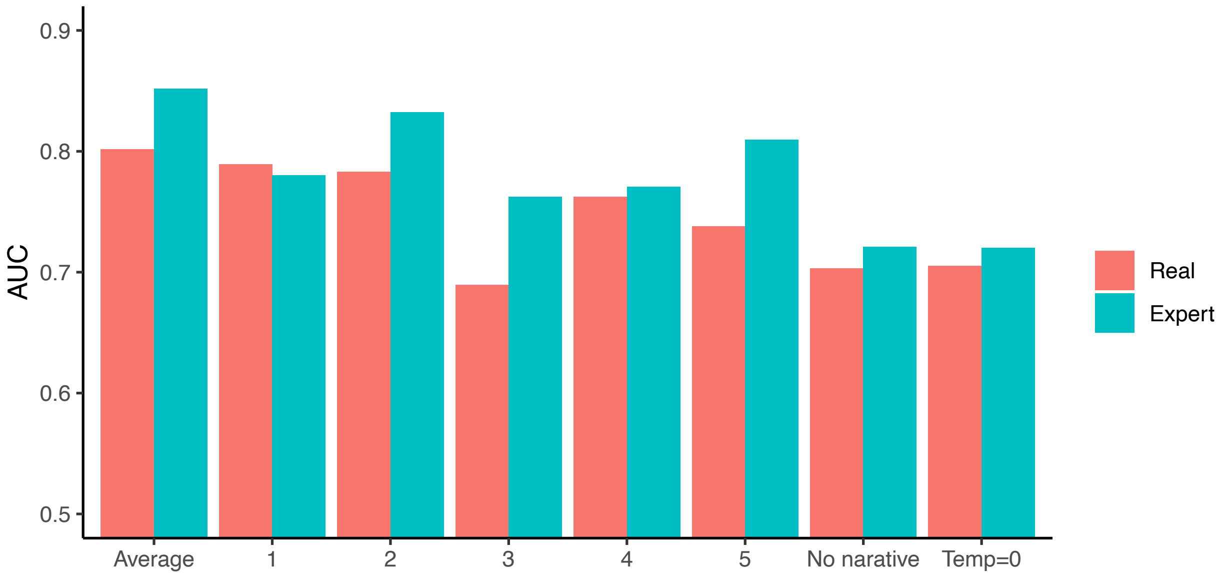


**Supplementary Figure 1. GPT-4 Assessments Performance.** Area under the curve (AUC) for GPT-4 decision to treatment scores of each of the individual submissions (1-5) and the average. Each individual submission is lower than the average. In addition, we submitted the cases without the clinical presentation narrative, which yielded lower AUC (no narrative). Similarly, lower AUC was observed when cases were submitted with temperature=0.
